# Beyond the Smoke: A Phenomenological Study of Health and Social Implications of Kush Use Among Sierra Leonean Youths

**DOI:** 10.1101/2024.04.24.24306083

**Authors:** Ronald Abu Bangura, Alhassan Mayei, Patrick Fatoma, Joseph Anderson Bunting-Graden, John Paul Kaisam, Rashid Ansumana

**Author notes:** Address for Correspondence: Rashid Ansumana, School of Public Health, College of Medical Sciences, Njala University, Bo Campus. These Authors contributed equally to the manuscript (Ronald Abu Bangura, Alhassan Mayei).

## Abstract

**Background:** The rising use of “Kush” among Sierra Leone’s youth is a public health concern. Kush, a concoction of Cannabis indica and synthetic substances, symbolizes the broader issue of drug misuse in low-income settings. This study explored the intricacies of Kush use among young Sierra Leoneans, highlighting the gravity of the crisis.

**Methods:** The research employed a phenomenological method, gathering insights through Focus Group Discussions (FGDs), Key Informant Interviews (KIIs), and In-Depth Interviews (IDIs) to understand the motivations and repercussions of Kush consumption. Selecting participants from diverse socio-demographic groups, the study included habitual users and those in vulnerable situations. Ten interviews were analyzed thematically, facilitated by NVivo software and Concept maps, to distill information.

**Results:** Findings identified numerous reasons for Kush use, from managing stress and trauma to seeking social belonging and economic necessity. Users experienced severe health issues, economic instability, and productivity loss. The effects varied by Kush strain, underscoring its heterogeneous nature. Despite a strong will to quit, influenced by health, family, and aspirations, youths faced hurdles like withdrawal symptoms, peer pressure, and scarce support systems.

**Conclusion:** Kush consumption in Sierra Leonean youths is entwined with socioeconomic and post-conflict factors, leading to significant adverse effects. Effective intervention requires a multifaceted strategy, focusing on underlying drug abuse causes and providing robust support systems. This study calls for tailored measures and policies that acknowledge the complex motivations behind Kush use and its impacts, aiming to foster an environment conducive to youth empowerment and health.

## INTRODUCTION

The surging public health dilemma posed by synthetic cannabinoids, exemplified by “Kush,” represents an escalating crisis in West Africa[1,2]. The designation “Kush” conjures images of the perilous Hindu Kush mountains, a metaphor for the formidable risks inherent to the drug. Kush, an artificial compound, is said to include cannabis, fentanyl, and tramadol, among other psychoactive elements, mirroring the dangerous qualities of the terrain after which it is named. The misuse of this synthetic cannabinoid by the youth of Sierra Leone has precipitated a significant uptick in morbidity and mortality, prompting the Government of Sierra Leone to declare a public health emergency on April 4, 2024, as reported by the British Broadcasting Corporation(BBC)[3]. This state of emergency highlights the compounding challenges faced by the population, particularly young men aged 18 to 25, amidst civil strifes, the aftereffects of the Ebola outbreak, and the socioeconomic disruptions engendered by the COVID-19 pandemic[2].

The synthetic characteristics of Kush, akin to other emerging psychoactive substances, present profound risks to public health. The diversity and often unsupervised nature of these compounds’ chemical structures contribute to unpredictable and severe health outcomes[4–8]. These outcomes are comparable to the detrimental effects of xylazine, colloquially termed the “Zombie” drug, which, when used in conjunction with fentanyl, has been associated with a troubling rise in overdose fatalities internationally[5,9,10].

The adverse consequences of Kush consumption extend to chronic cognitive impairments, anxiety, and paranoia[6]. Echoing the effects of xylazine, the proliferation of Kush correlates with societal issues such as heightened rates of violent crime, increasing substance misuse, disrupted family structures, and diminished youth productivity. Consequently, Kush is exacerbating the already fragile socio-economic fabric of Sierra Leone and other West African countries such as Liberia[11], Guinea and Ghana[12] and Nigeria[13] having the same drug problems, hampering their economic recovery and development[5].

There exists a substantial lacuna in the literature concerning the intricacies of Kush and its ramifications, particularly in African countries grappling with socio-economic instability. This study intends to delve into the phenomenology of Kush usage, endeavoring to fill the knowledge gap and guide effective public health strategies.

## METHODS

### Research Design

We used a phenomenological technique, a type of qualitative research design to explore the multiple perspectives and complex perceptions that young people relate to the use of kush. To understand the fundamental reasons behind Kush usage, its socioeconomic drivers, and the subsequent effects on health and social life, the study focused on the lived experiences and subjective perspectives of users.

This approach was chosen for its strength in uncovering the depth, complexity, and contextual factors influencing participants’ lived experiences and behaviours related to Kush use. Focus group Discussions (FDGs), Key informant interviews, and In-Depth interviews were conducted to facilitate rich, interactive discussions among participants, allowing for the emergence of diverse perspectives and insights into the social dynamics of substance use. Discussion in groups usually elicits new information and allows members of the group to question one another on the information they are providing. This focus group process often uncovers additional information not discovered in the individual interviews.

### Participant Selection

We purposively selected participants between the ages of 18 and 30 years who self-identified as current or former Kush users. The target population consisted of people who use kush frequently and spend a lot of time in secluded areas including ghettos, car washes, parking lots, and garages in Bo. This group was diverse, including motorbike riders, youths engaged in car or motorbike washing, kush or tramadol sellers, and auto-mechanics, representing a wide range of socio-demographic backgrounds. This age group was purposely chosen to cover a wide array of the youth population that might be impacted by substance use. Participant recruitment was aided by local community leaders and youth organizations in Sierra Leone to avoid ethical breaches and to build trust with the participants. We conducted seven Focus Group Discussions (FGDs), two Key Informant Interviews (KIIs), and one In-Depth Interview (IDI), totaling ten interviews. Each FGD included 6-8 participants, which yielded a richly diverse sample that reflected varied socio-economic statuses and experiences with kush use and motivations.

### Data Collection

Focus group interviews were held in confined venues within communities, facilitated by trained moderators experienced in qualitative research and sensitive to the nuances of discussing substance use. Each session lasted approximately 90 minutes and was structured around a semi-structured interview guide. The guide included open-ended questions designed to elicit detailed responses on participants’ motivations for using kush, their experiences with different types of kush, and the perceived impacts on their health, social life, and economic well-being. To ensure accuracy and facilitate analysis, all focus group sessions were audio-recorded, with participants’ informed consent, and subsequently transcribed verbatim.

### Ethical Considerations

The study was conducted keeping ethical guidelines in mind for research involving human subjects. Before interviews, all participants were informed about the purpose of the study, the voluntary nature of their involvement, the right to decline or refuse to answer any question that they felt uncomfortable answering, and their right to withdraw at any point without consequence. Informed consent was obtained from each participant, ensuring confidentiality and anonymity in the handling of their data. The study protocol, including recruitment strategies and consent procedures, received approval from the Njala University ethical review board.

### Data Analysis

Thematic analysis, which is particularly useful for identifying, assessing, and uncovering patterns within qualitative data, was applied to the focus group discussion transcriptions of the data. Following Braun and Clarke’s six-phase approach[14], this analysis began with familiarising with the data and proceeded to generate preliminary codes, search for themes, review themes, define and name themes, and finally compile the report. NVivo, a programme for analysing qualitative data, was used to help with the coding and topic creation. Using a concept map, important themes that emerged from the analysis were visualised. The approach was incremental, with themes constantly being improved upon as new information emerged from the data.

### Rigor and Validity

We used multiple techniques to guarantee validity and rigour in the qualitative analysis. Peer debriefing sessions among the research team to debate and improve the coding and theme development procedures were among them, as was the triangulation of data sources through several focus groups, and member checking with participants to validate interpreted findings. Additionally, the researchers engaged in reflexivity by recognising their limitations and considering how their personal experiences and opinions might have affected the analysis.

### Results and Insights

The thematic analysis of focus group interviews with Sierra Leonean youths provided good knowledge on the motivations for, and effects of, Kush use. This combined results section integrates findings from initial and expanded analyses, merging the complex motivations behind Kush consumption and its multifaceted impacts on users’ lives, supported by direct quotations from participants.

### Motivations for Kush Use

Participants mentioned a range of motivations that make them take kush, with stress and personal challenges being paramount. The use of Kush as a coping mechanism for dealing with the aftermath of Sierra Leone’s civil war, economic hardships, and personal losses was prominently mentioned. “*After my parents died, I just couldn’t find any peace until I started hanging out with friends who introduced me to Kush. It kinda helps me forget*,” revealed Participant 4, Male, 21-25 years old, underscoring the psychological escape Kush provides from life’s adversities.

Peer influence and the desire for social belonging also emerged as significant factors. “*Everyone in my circle was doing it. Not joining in felt like I was on the outside*,” Participant 8, Female, 18-20 years old, highlighted the social pressures and the role of Kush in fostering a sense of community among peers. Economic and work-related pressures further compounded the reasons for Kush use, especially among those engaged in manual labour or facing unemployment. “*It gives you the energy to keep going, even when your body wants to quit. It’s either that or lose a day’s pay*” Participant 11, Male, 21-25 years old, indicated the reliance on Kush for enduring physically demanding tasks(Fig. 1).

**Figure 1:**
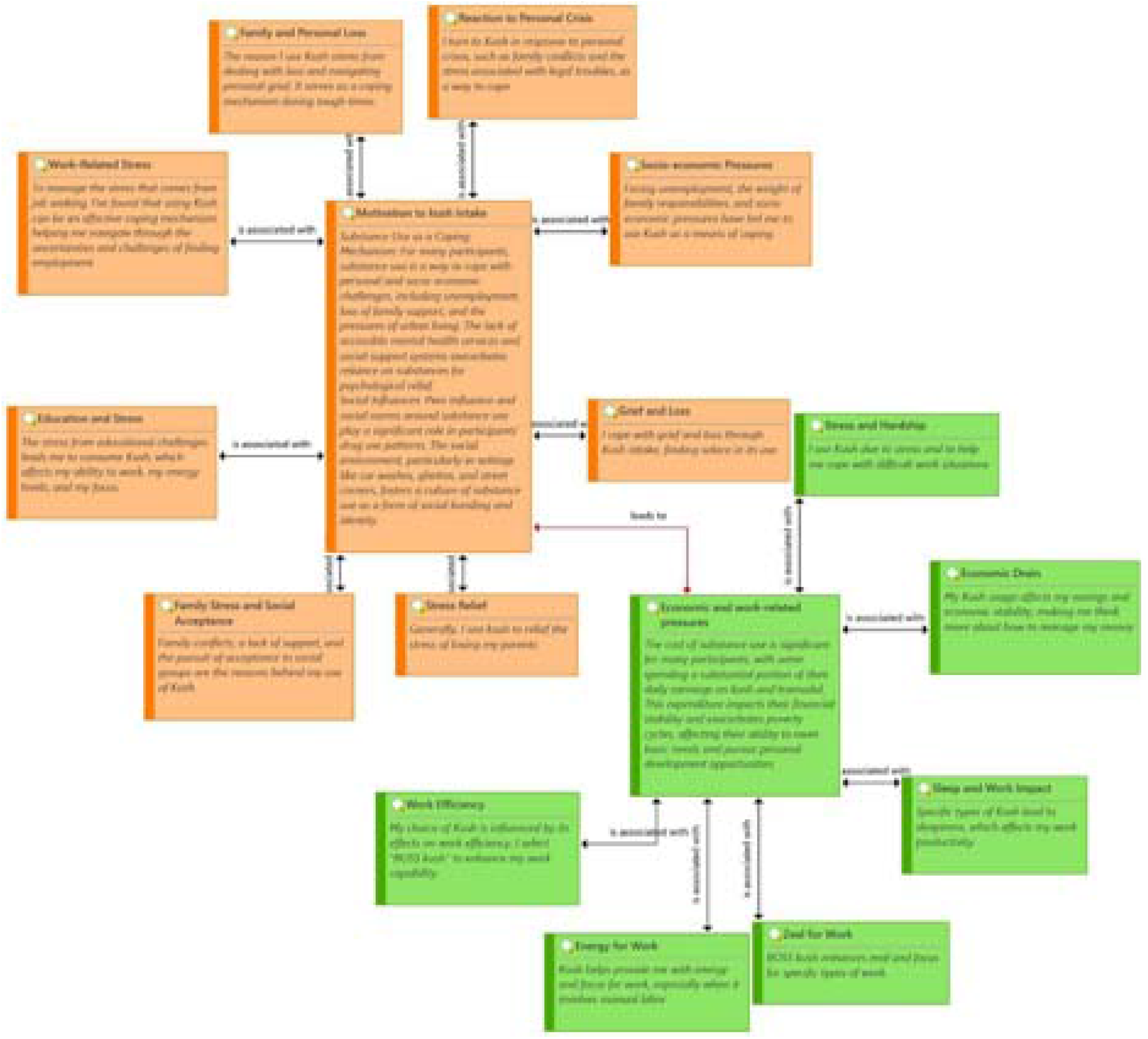
The Complex Interplay Between Stress, Kush Use, and Socio-Economic Outcomes.

### Impacts of Kush Use

The impacts of kush use spanned several domains, including physical health, psychological well-being, social relationships, and economic stability.

#### Physical Health

Participants reported both positive and negative physical effects of kush use. Some highlighted its energy-boosting benefits, while others experienced adverse outcomes. “*I’ve developed this cough that just won’t go away. I know it’s from the smoking*” Participant 9, Female, 18-20 years old, shared, pointing to the respiratory issues associated with chronic Kush smoking.

#### Psychological Well-being

Kush use affected participants’ mental health, with reports of cognitive impairment and emotional instability. “*I find it hard to remember things these days, like my mind is always foggy*,” Participant 7, Male, 21-25 years old, described the cognitive fog linked to regular use.

#### Social Relationships

Kush use had a profound impact on users’ social dynamics, often straining familial ties and friendships. “*My family doesn’t look at me the same way anymore. They think I’m throwing my life away*” Participant 13, Male, 18-20 years old, illustrated the social repercussions of perceived substance misuse.

#### Economic Stability

The financial burden of sustaining kush use was a recurrent theme, with many participants lamenting the significant portion of their income spent on kush. “*Most of my money goes to buying kush. It’s hard to save anything*” Participant 25, Male, 26-30 years old, highlighted the economic implications of habitual kush use.

### Various Types of Kush Consumed

Youths described several experiences that vary depending on the type of kush they use when speaking about their usage of different kush strains(Fig. 2). Their experiences illustrate how various kush varieties influence them and the decisions they make according to their kush-related goals. A 21-25-year-old young man mentioned “Boss Kush” and how it supports his increased productivity and alertness. “*After using ‘Boss kush,’ I feel as though I can work for hours without feeling fatigued*,” they remarked. When I have a lot on my plate or need to study a lot, it comes in incredibly handy. Many who require more energy and concentrate on work or other tasks choose this strain of kush.

**Figure 2:**
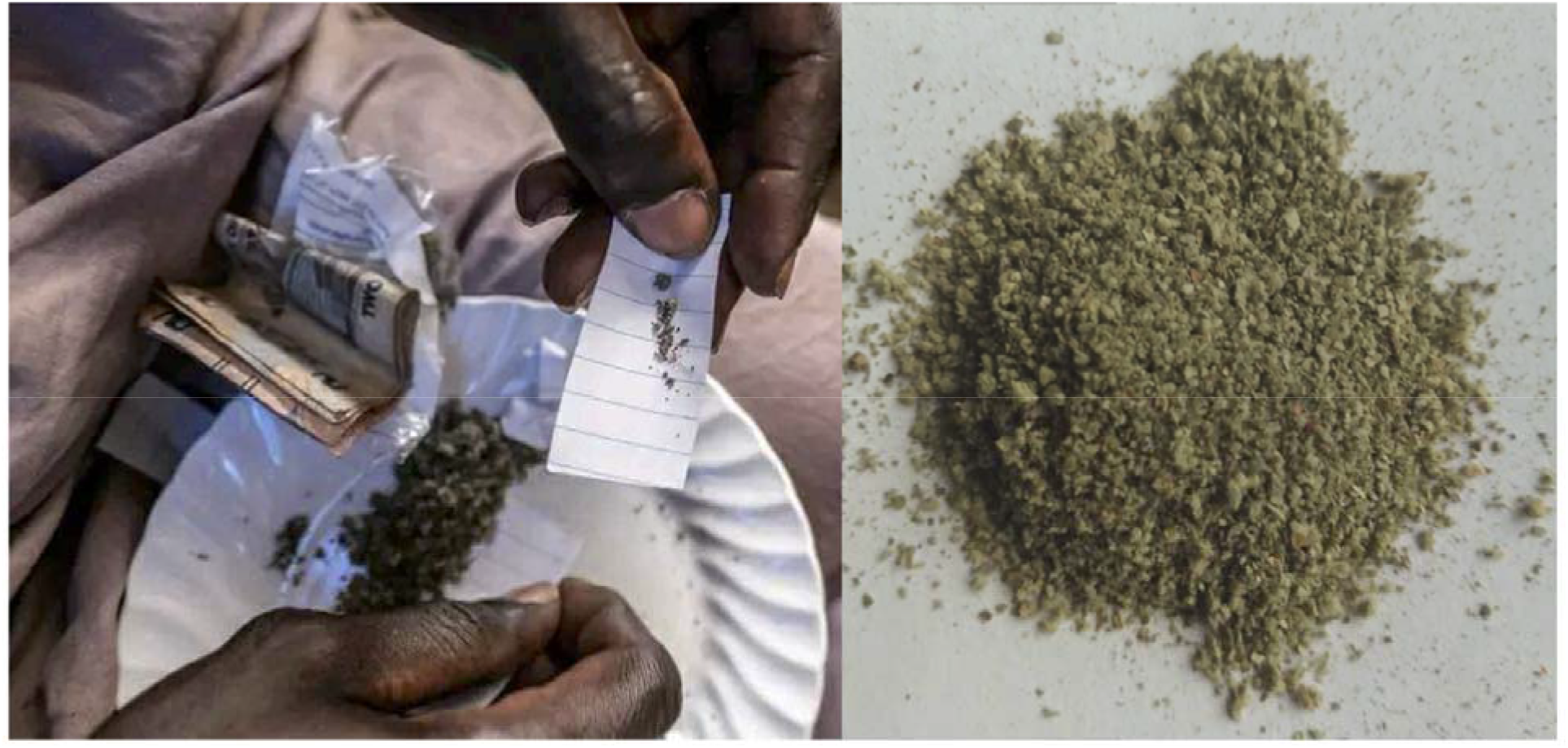
Some Examples of Kush

A 21-25-year-old individual also mentioned experiencing a powerful high that sharpens their senses after using “Jagaban kush”. “*The feeling you get from ‘Jagaban kush’ is really strong; it makes everything feel more intense*.” This indicates that certain young individuals select this strain of kush in search of a potent high (Figure 1).

However, one 18-20-year-olds stated that they prefer “Gentle kush” since it makes them relax and have better sleep. They mentioned, “*‘Gentle kush’ helps me relax and get a good night’s sleep after a hectic day*.” *When I’m tired, it’s my favourite*.” It would appear from this that some young people take kush for energy, whereas others use it for relaxation and sleep. There are significant risks associated with certain forms of kush. A 26-31-year-old warned about the hazards of “Tramadol kush,” adding, “*Seeing someone vomit blood after using ‘Tramadol kush’ is alarming*”. It shows how harmful it may be. This highlights the serious health concerns associated with taking specific types of kush, demonstrating that it is not always harmless.

These youth experiences demonstrate the various reasons individuals use kush, ranging from wanting more energy to needing help sleeping, as well as the major risks associated with some varieties. By sharing their experiences, we gain a deeper understanding of why young people use kush and the effects it has on them. This information is important for helping everyone understand the complex reasons behind kush use and the need to be careful about its dangers.

### Desires to Stop Kush Use: Insights from Sierra Leonean Youths

The thematic analysis of focus group discussions revealed a significant inclination among participants towards ceasing Kush use, driven by various factors and accompanied by diverse strategies and challenges. Participants shared their motivations for wanting to quit, the method they consider or have attempted to employ in cessation efforts, and the obstacles they face in this endeavour. Their narratives provide a comprehensive view of the journey towards cessation.

### Motivations for Cessation

A prevalent theme was the desire for improved physical health, with participants expressing concerns over the negative health impacts associated with prolonged Kush use. “*I started feeling this constant fatigue, and it scared me. I knew then I needed to stop for the sake of my health*” a participant recounted highlighting health as a primary motivator for cessation. (Participant 18, Male, 21-25 years old)(Figure 3).

**Figure 3:**
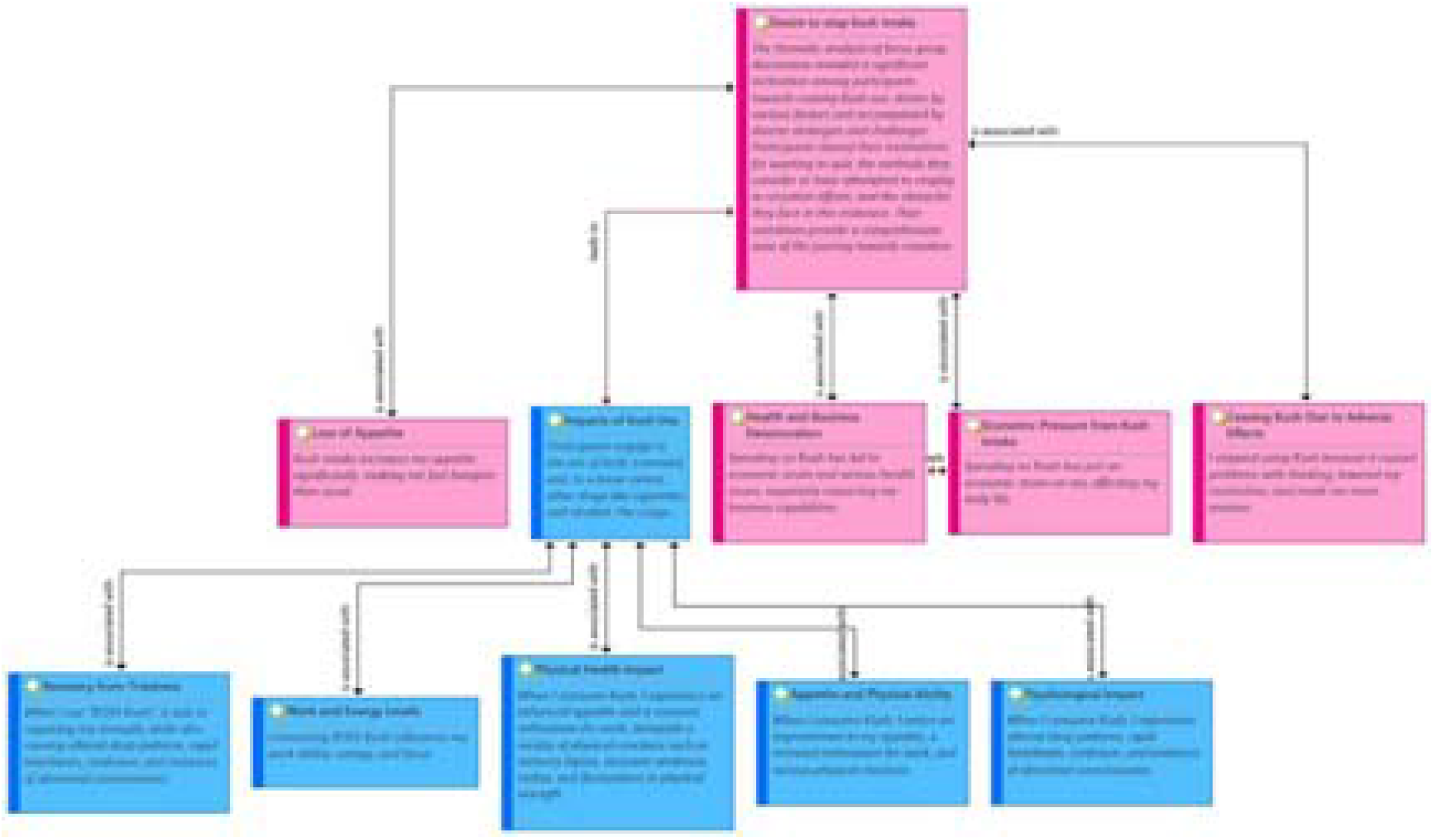
Understanding the Multifaceted Effects of Kush Use and the Journey Towards Cessation

Family and future aspirations also played a crucial role in participants’ desire to quit Kush. The impact of substance use on familial relationships and future opportunities was a significant concern. “*My sister told me she believed in me, that I could do more with my life*… *It made me want to be better*,” shared another participant (Participant 21, Female, 18-20 years old), emphasizing the influence of family support and personal goals on the decision to quit. Strategies for Cessation.

Social support and positive peer influence emerged as pivotal in the cessation process. Participants discussed the importance of being surrounded by friends who support their decision to quit or who are also in the process of quitting. “*Having a buddy who’s also trying to quit helped a lot. We keep each other in check*,” one participant noted (Participant 25, Male, 21-25 years old), illustrating the value of mutual support.

The need for and scarcity of professional support services, including rehabilitation and counseling, were frequently mentioned. Despite the desire for professional help, barriers such as limited access and stigma were significant obstacles. “*I wanted to get help, but there’s not much out there. And there’s shame in asking*,” a participant lamented (Participant 27, Male, 21-25 years old), pointing to the challenges in accessing support.

Engaging in alternative activities to occupy time and redirect focus from Kush was another strategy discussed. Participants highlighted the role of hobbies, sports, and vocational training in their efforts to quit. “*I started playing football again, and it’s been great*… *Keeps me busy, and I don’t think about smoking as much*,” said a participant (Participant 30, Male, 21-25 years old), underscoring the importance of finding new interests.

### Challenges in Cessation

Despite the strong desire and various strategies to quit Kush, participants faced numerous challenges. The physical and psychological withdrawal symptoms, social pressure from peers still using Kush, and the lack of alternative coping mechanisms for stress and trauma were significant hurdles. Moreover, the absence of accessible and effective support services compounded the difficulty of cessation efforts.

### The Use of Bones from Cemeteries as a Mixture for Kush

Bones were reported dug from cemeteries and ground to dust and then mixed with other elements in the kush mixture. The bones were obtained only from Christian graves and bones of animals in butcher markets were not used. In Sierra Leone, Christians are normally preserved and not buried on the same day of death, the chemicals used as embalming fluid include formaldehyde, which is a major constituent of the mixtures used in kush and not normally in high supply in Sierra Leone.

## Discussion

The findings from the focus group interviews with Sierra Leonean youths offer profound insights into the multidimensional nature of Kush use, its motivations, impacts, and the desires surrounding cessation. These insights not only illuminate the personal and societal dynamics of Kush use in Sierra Leone but also contribute to the broader discourse on youth substance use in post-conflict and socio-economically challenged settings.

The motivations for Kush use—ranging from coping with stress and trauma to social integration and economic survival—reflect a complex interplay of psychological, social, and economic factors. This complexity underscores the inadequacy of viewing substance use merely through the lens of individual choice or moral failing. Instead, it highlights the necessity of understanding substance use within the broader context of users’ lives, including the lingering effects of Sierra Leone’s traumatic events such as the Ebola Outbreak and current socio-economic challenges.

The study identified an intricate interplay of social, cultural, and economic factors that affect kush consumption. The results imply that people use kush as a means of coping to try to lessen different kinds of stress. According to the findings, stress due to unemployment is a major factor in Kush use. Many claim that using Kush helps them stay productive and handle the demands of their work. Another factor is educational stress, as young people and students use Kush to deal with the pressures of unemployment and school, which impairs their focus, energy, and ability to concentrate.

Personal problems, including the loss of a family member and conflicts within the family, are important drivers of Kush usage. Participants said that Kush provides a way to escape from the difficulties of family life and aids in navigating complicated emotional landscapes. Additionally, some turn to Kush as a coping mechanism for the socioeconomic stresses in their lives, such as financial obligations and unemployment. In terms of social interactions, Kush is utilized to strengthen bonds in some communal contexts where it is widely used. Some people use it as a social glue, helping those in organisations where usage is a prevalent bond and be accepted. The study made clear the long-term economic effects of regular Kush usage, even about any apparent short-term benefits. The cost of keeping up a Kush habit was high; some people had to set aside a sizable amount of their money to maintain their usage, which made their already pre-existing financial concerns worse and maybe put them in a vicious cycle of poverty. Although some users reported that Kush increased their energy and productivity at work, others reported that it had a detrimental effect on their sleep and overall productivity. This shows that there may be a complicated connection between the short-term advantages of improved focus and the long-term harm to one’s health and financial security.

As identified by the participants, Kush use has multifaceted effects that cut across physical health worries, psychological and cognitive changes, social relationship straining, and economic hardships, among others. These multiple effects[15–17] call for comprehensive interventions that cover not just the health-related aspects of substance use but also its social[18] and economic dimensions. Analysis of the focus group discussions revealed a strong desire among the participants to quit Kush, driven by the multifactorial effects and the need to avoid any chance of the substance’s ill effects. Specifically, the participants reported their reasons for wanting to quit, the approaches they use or would consider, and the challenges that they face in such pursuit. The desire voiced out by most participants and the approaches identified or considered are indicative of a substantive recognition of Kush’s ill effects and a craving for change. For example, most of the participants expressed increased appetite following Kush usage, with one saying *“It makes me feel hungrier than ever”*. Others cited “*BOSS Kush*” as a potent mixture affecting their ability to work, their energy levels, and their concentration, as well as aiding in fatigue recovery. However, these identified advantages are accompanied by detrimental physical reactions such as memory lapses, bodily weakness, rashes, and fluctuating physical energy. Also, the psychological effects of Kush were pronounced as identified by users who reported alterations in sleep patterns, rapid heartbeats, confusion, and abnormal consciousness. These psychological effects outline an alarming aspect of Kush’s effect on the users. The identified outcomes of kush include economic repercussions as most of the users identified kush expenditure as a major economic constraint, affecting daily activities and businesses. Usually, the economic burden that is associated with the substance worsens the health concerns, creating adverse personal and professional effects. Ultimately, due to the identified multifactorial effects,, some of the participants have withdrawn from the usage of kush following severe effects that they experienced and shared. Such effects included thinking problems, lowered motivation, and increased levels of anxiety. Such identified factors outline an essential component of substance use on the one hand and the dependence on use and the severe effects that drive the action on the other. Particularly for the participants, the journey to quitting kush is dotted with challenges but also defined by a substantive recognition of the multiple effects of the substance.

However, the limitations concerning the withdrawal symptoms, social consequences, and the design of existing support networks reveal considerable drawbacks in the state of assistance for drug users in Sierra Leone. There is clearly and urgent need for a multisectoral, community-centred, stakeholder-engaged, collaborative intervention[19,20].

The results of this study carry several implications for further research and practice, as well as policy. Among the most critical, there is a need to develop and ensure access to a comprehensive support system for young people who wish to stop drugs. Such a system should include easily accessible community support pathways, as well as help lines and mental health and rehabilitation services in addition to various educational interventions addressing the root causes of drug use. Additionally, policies responding to youth substance use in Sierra Leone or elsewhere in the world, and similar contexts, should be holistic and strive to address the underlying socio-economic conditions promoting drug consumption by creating educational, employment, and recreational opportunities. Future research might take the form of elaborating on the complexities of reasons for and outcomes of using substances. Specifically, future studies might include gender analysis, an examination of new digital media, and an evaluation of the stability of cessation support. Longitudinal studies to evaluate the long-term consequences of substance abuse would also be interesting to consider in the future.

Several limitations to this study could affect its generalizability and the robustness of the results. The most important limitation is the use of purposive sampling and a relatively small sample size, which could limit the diversity of perspectives and result in selection biases. The urban background of all the participants also makes it impossible to generalize the conclusions across the rest of the Sierra Leonean youth population. Secondly, the study’s qualitative and self-reported design might introduce sources of bias, such as recall bias, or social desirability bias, which could affect the truthfulness of the reported reasons and effects of using kush. The cross-sectional design does not allow for assessing the changes over time or the long-term implications of kush use. Lastly, the lack of quantitative data validating and extending the findings from the interviews also weakens the conclusions.

Notwithstanding, we share the perspectives reported in this article to extend the knowledge of kush and its multifaceted effects on youths.

## Conclusion

The analysis of kush use among young people in Sierra Leone uncovers a story deeply embedded in the country’s socio-economic and post-conflict fabric. While the academic motivations behind kush use are diverse, they are all linked to stress, trauma, and finding one’s place and existence. Similar complexities are demonstrated in how they are affected and provided to cease using substances. To adequately address kush use among young people in this population, a compassionate, comprehensive approach that acknowledges the variety of factors influencing substance use is required. The intervention should be informed by the socio-economic context of young people’s life, provide support beyond cessation, and aim at overall well-being and empowerment. This paper addressed critical areas of need and potential pathways for assisting young Sierra Leoneans in managing kush use. It requires a collaborative effort from policymakers, community leaders, health practitioners, and academics to create a future where young people can flourish without substance constraints.

Moreover. the story of kush use among the young generation in Sierra Leone is loud and widespread, but more than that, it represents the narrative of the neighboring West African countries. Nigeria[13], Ghana, and Liberia[11,12,21] are also among the states struggling with the socio-economic challenge of substance use, and, perhaps, the history of conflict fueling the phenomenon. The factors motivating the youth in these nations to resort to substances such as kush appear to be consistently linked to underlying issues of poverty, lack of education, and the enduring effects of warfare or civil unrest. The countries of West Africa must devise strategies that are attuned to their cultural and regional contexts, addressing not only the cessation of kush but also providing a comprehensive array of socioeconomic and psychological support. These strategies must take into account the societal and cultural dimensions of substance use. Regional health officials, custodians of tradition, healthcare providers, and researchers ought to prioritize sensitivity to these issues, thus forging pathways for the youth to seek fulfillment and success without resorting to drugs. Collaboration and the sharing of successful strategies among these neighboring countries could be invaluable, fostering a unified commitment to nurturing a resilient and thriving West African youth population.

There is a need for further studies with additional methodological rigor through quantitative analyses and chemical analyses for clarity on the toxicity of the different brands of kush and the potentially varied implications on human health.

## Data Availability

All data produced in the present study are available upon reasonable request.

https://1drv.ms/w/s!AvaQPIzRgvoecKcCA-KcID-kcBE?e=LhMsAQ

## Data availability statement

Data are available upon request. Data can be accessed and downloaded from the following URL: https://1drv.ms/w/s!AvaQPIzRgvoea76c2pTr8MbpqF0?e=S2on4G

## Ethics statements

Patient consent for publication: Not required.

## Ethics approval

This study was approved by the Njala University Institutional Review Board

## Acknowledgments

We acknowledge the faculty of the School of Public Health, Njala University, the research subjects, who shared heir life experiences with us and the West African One Health Consortium for providing the space for capacity building which culminated to this piece.

X: @ansumanaR, @banguraronaldA

## Contributors

RAB and RA conceived the study. RAB, RA, AM, PF, designed the study. RAB, AM, PF, RA, were involved in data collection and analyses. RAB, AM, RA, PF, JPK and JAB were involved in drafting and critically reviewing the manuscript. All authors read and approved the final manuscript

